# Sex-specific association of cardiovascular drug doses with adverse outcomes in atrial fibrillation

**DOI:** 10.1101/2024.04.20.24306039

**Authors:** Jeanne Moor, Michael Kühne, Giorgio Moschovitis, Richard Kobza, Seraina Netzer, Angelo Auricchio, Jürg Hans Beer, Leo H. Bonati, Tobias Reichlin, David Conen, Stefan Osswald, Nicolas Rodondi, Carole Clair, Christine Baumgartner, Carole Elodie Aubert, the BEAT-AF and Swiss-AF investigators

## Abstract

**Objectives:** Women with heart failure (HF) with reduced ejection fraction receiving submaximal doses of beta-blockers and renin-angiotensin system (RAS) inhibitors have a lower risk of mortality or hospitalizations for heart failure. However, optimal doses of beta-blockers or RAS inhibitors in women with atrial fibrillation (AF) with and without HF are unclear. We investigated sex-specific associations of beta-blocker and RAS inhibitor doses with cardiovascular outcomes in patients with AF with and without HF.

**Methods:** We used data from the prospective BEAT-AF and Swiss-AF cohorts on patients with AF. The outcome was major adverse cardiovascular events (MACE), including death, myocardial infarction, stroke, systemic embolization, and HF-related hospitalization. Predictors of interest were spline (primary analysis) or quartiles (secondary analysis) of beta-blocker or RAS inhibitor dose in percent of the maximum dose (reference), in interaction with sex. Cox models were adjusted for demographics, comorbidities and co-medication.

**Results:** Among 3,961 patients (28% women), MACE occurred in 1,113 (28%) patients over 5-year median follow-up. Distributions of RAS inhibitor and beta-blocker doses were similar in women and men. Cox models revealed no association between beta-blocker dose or RAS inhibitor dose and MACE. In a subgroup of patients with AF and HF, the lowest hazard of MACE was observed in women prescribed 100% of RAS inhibitor dose. However, there was no association between RAS dose quartiles and MACE.

**Conclusions:** In these two cohorts of patients with AF, doses of beta-blockers and RAS inhibitors did not differ by sex and were overall not associated with MACE.

**What is already known on the subject:** Sex-specific analyses of beta-blocker and renin angiotensin system (RAS) inhibitor doses in patients with heart failure with reduced ejection fraction have revealed a lower hazard of death or heart failure-related hospitalisation in women receiving low doses compared to maximum doses.

The pathophysiology and pharmacotherapy of atrial fibrillation show sex differences, but the potential sex-specific associations of different drug doses with cardiovascular outcomes are unknown in this population.

**What this study adds:** This study identifies no associations between beta-blocker doses and major adverse cardiovascular events in patients with atrial fibrillation.

**How this study might affect research, practice or policy:** The findings of the present study reassure that the recommended maximum doses of beta-blockers and RAS inhibitors appeared safe among patients of both sexes with atrial fibrillation.

## INTRODUCTION

Atrial fibrillation (AF) is currently the most prevalent arrhythmia, and its incidence is increasing while survival of patients with AF has not improved in recent years (1). Among patients with AF, up to 77% suffer from concomitant heart failure (HF) which is a predominant cause of death (2). In patients with HF with reduced ejection fraction (HFrEF), positive effects on clinical outcomes are established for several drug classes including beta-blockers and renin angiotensin system (RAS) inhibitors. However, the evidence for a benefit of beta-blockers in patients with AF only is limited to ventricular rate control and for prevention of symptomatic AF (3–5). RAS inhibitors may prevent incident and recurrent episodes of AF in some populations, for example AF recurrence in Asian but not in European or American populations (6,7).

Sex differences are increasingly recognized in AF. The age-adjusted prevalence of AF is lower in female compared to male sex, but female sex is associated with a lower quality of life and a higher hazard of complications of AF such as stroke and cardiovascular death (8,9). In addition, sex differences exist in treatment allocation in patients with AF, with a higher proportion of men receiving electric cardioversion, radiofrequency ablation or pharmacotherapy compared to women (10). Cardiovascular pharmacokinetics also differ between the sexes, with beta-blockers causing higher peak concentrations in women (11), and angiotensin converting enzyme (ACE) inhibitors carrying a larger distribution volume and residency time in women (12). Further, the risk of adverse effects of ACE inhibitors is higher in women compared to men (13). However, the potential impact of these sex differences in pharmacokinetics on cardiovascular outcomes is unclear.

Recent data show that optimal doses of beta-blocker and RAS inhibitors may differ between women and men. In two European and Asian populations with HFrEF, women receiving submaximal doses of RAS inhibitors or beta-blockers showed a lower hazard of mortality or cardiovascular hospitalizations compared to women receiving maximal doses (14). Similarly, in Dutch outpatient clinics, a RAS inhibitor dose <50% was associated with lower mortality in women but not in men with HFrEF (15). However, the sex-specific optimal doses of beta-blockers in patients with AF and doses of RAS inhibitors in patients with both HF and AF are unclear. In addition, despite the lack of a clear benefit of RAS inhibitors in patients with AF only, potential dose-dependent and sex-specific benefits or harms could exist.

Our goal was thus to assess sex-specific associations between beta-blocker or RAS inhibitor dose and major adverse cardiovascular events (MACE) in (i) patients with AF, and (ii) in patients with AF and HF.

## METHODS

### Study design and population

The current study is a post-hoc analysis of patients with AF from two prospective cohorts that were developed to determine cardiovascular and neurological outcomes in patients with AF: the Swiss Atrial Fibrillation (Swiss-AF) Cohort Study and Basel Atrial Fibrillation (BEAT-AF) Cohort Study (16,17). Swiss-AF and BEAT-AF are prospective multicentre cohort studies across 14 and seven Swiss centres, respectively (17). Both studies enrolled adult patients from inpatient and outpatient clinics, BEAT-AF between January 2010 and April 2014 and Swiss-AF between March 2014 and August 2017 (16,17). Enrolment required at least one documented episode of AF on an electrocardiogram and written informed consent. The Swiss-AF cohort excluded patients with secondary forms of AF (e.g. onset after surgery) and with any acute illness within the last 4 weeks (17). The BEAT-AF cohort had no major exclusion criteria (16). All study sites had received approval by the local ethics committee, and the studies were conducted with adherence to the principles of the Helsinki Declaration.

### Outcomes

The primary outcome was time until the first event of a composite of MACE defined as all-cause death, myocardial infarction, coronary revascularization, stroke, and hospitalization due to HF (18). Secondary outcomes were the first event of individual components of the primary outcomes. All outcomes, except coronary revascularization, were predefined in the protocol of the cohort studies (16,17), ascertained by annual visits and adjudicated by a blinded committee of clinical experts.

### Exposures

Doses of beta-blockers, ACE inhibitors and angiotensin receptor blockers and sex were the main variables of interest.

Drugs and their respective doses were identified from digital study records of medication lists at baseline visit using a systematic automated text search for all compounds under the categories of beta-blockers, ACE inhibitors and angiotensin receptor blockers (Supplemental Table 1). Relative daily beta-blocker drug doses were calculated as percent of maximum doses according to the 2020 European Society of Cardiology (ESC) guidelines for AF (4). RAS inhibitor doses were calculated as percent of maximum doses in ESC guidelines for HF (3). Wherever the two guidelines articles did not include an individual beta-blocker or RAS inhibitor compound, equivalence doses were determined using a web-based calculator (19). Further, demographic information, comorbidities and further medication data were collected by investigators during study visits using electronic questionnaires and were included in the present analysis. Body surface area was calculated using the formula of Du Bois (20).

### Statistical analysis

Based on previous data, we calculated that 509 events would be needed to obtain 80% power (alpha 0.05) to detect a relative hazard for mortality or heart failure-related hospitalization of 0.78 in women treated with submaximal doses of RAS inhibitors and 900 events to detect a relative hazard of 0.84 in women treated with submaximal doses of beta-blockers compared to women treated with the maximal doses, respectively (14,21). Continuous variables are reported as median (interquartile range [IQR]) and categorical variables as frequency (percentage). We analysed the overall population and the subset of the patients with a history of HF at baseline according to a pre-specified analysis plan. As a first analysis strategy, beta-blocker or RAS inhibitor drug doses in percent were used as continuous variable (14,22). As a secondary analysis strategy, beta-blocker and RAS inhibitor doses were categorized in five groups containing those with 0% (those without the drug) and the quartiles of the subset prescribed between 1 to 100%. Participants with >100% of maximum allowed daily dose of beta-blockers (n=5) or RAS inhibitors (n=22) were excluded from the respective analyses. The hazard of primary and secondary outcomes was calculated using Cox proportional hazards models. For the primary analysis strategy, the restricted cubic spline of drug dose was used as nonlinear predictor in Cox models. For the secondary analysis strategy, drug dose categories were included in Cox models. The top quartile or 100% of drug dose (the maximum beta-blocker or RAS inhibitor dose) in men was set as reference, respectively. Additional variables of interest included in the models were sex, and the interaction between drug dose and sex. Models with and without the interaction term were compared using Likelihood Ratio tests, with p<0.05 considered as significant. Models were adjusted for covariates identified from the literature as potential confounders (14,23), including age, body surface area (BSA), current smoking status, regular physical activity, history of diabetes, chronic kidney disease, history of coronary artery disease, heart rate, hypertension, history of stroke and/or transient ischaemic attack, oral anticoagulation, antiplatelet therapy, antiarrhythmics, COPD or asthma, pre-existing HF, and the dose percentage of the other drug class (RAS inhibitors for the beta-blocker models, and vice versa) at baseline. Patients with loss of follow-up were censored at the last completed visit or recorded event. For secondary outcomes not including all-cause mortality, patients were censored by loss of follow-up and additionally at death. Missing data among model covariates underwent multiple imputation. Pre-specified sensitivity analyses were the adjustment for left ventricular ejection fraction (LVEF) in the subset with available echocardiography data, once as a continuous and once as categorical variable (cut-offs: <40%, 40-49%, 50% and above); the normalization of drug dose through division by BMI, BSA or body weight or the use of BMI instead of BSA in the models because of their discordance at extreme values (24); excluding those not prescribed a drug (beta-blockers or RAS inhibitors) in the respective models; and inclusion of AF-specific parameters (device, cardioversion, AF type, AF duration) as potential confounders in the models. As posthoc sensitivity analyses, the analysis was stratified for the different treatment indications, and inverse probability weighting was used to balance patient characteristics. All analyses were performed with RStudio 2023.06.1.

## RESULTS

### Demographic and clinical characteristics

Among the 3961 participants of the Swiss-AF and BEAT-AF cohorts, 28% were women, median age was 72 (IQR: 66, 78) years, and the most frequent type of AF was paroxysmal (49%). Women were of similar age to men, median BSA was 1.76m^2^ (IQR: 1.65, 1.88) in women vs 2.01m^2^ (IQR: 1.89, 2.13) in men. Common comorbidities included arterial hypertension (69%), coronary artery disease (27%), diabetes mellitus (16%) and chronic kidney disease (19%). Prevalence of coronary artery disease was 15% in women and 31% in men. Median LVEF was 60% (IQR: 55%, 65%) in women vs 55% (IQR: 47%, 60%) in men at baseline in the subset with available echocardiography data. Full demographic and clinical characteristics of the overall population are shown in Table 1. Characteristics of the twenty-five percent of men and 21% of women with a history of HF at baseline are shown in Supplemental Table 2. MACE occurred in 815 (29%) men and 308 (28%) women over a median follow-up of 4.7 (IQR: 3.0; 6.0) years. Secondary outcomes included 632 deaths, stroke in 189, myocardial infarction in 137, hospitalization due to HF in 584 and systemic embolism in 16 participants.

**Table 1:** Baseline characteristics.

| **Table 1: Baseline characteristics** | **Overall**<br>**n = 3,961** | **Men**<br>**n = 2,844** | **Women**<br>**n = 1,117** |
| --- | --- | --- | --- |
| **Age (years)** | 72 (66, 78) | 72 (66, 78) | 74 (68, 80) |
| **BMI (kg/m<sup>2</sup>)** | 26.8 (24.2, 30.0) | 26.9 (24.6, 29.9) | 26.0 (23.0, 30.4) |
| **BSA (m<sup>2</sup>)** | 1.95 (1.81, 2.09) | 2.01 (1.89, 2.13) | 1.76 (1.65, 1.88) |
| **Heart rate (min<sup>-1</sup>)** | 67 (59, 78) | 66 (58, 77) | 68 (60, 79) |
| **Smoking status** |  |  |  |
| No | 1,739 (44%) | 1,091 (38%) | 648 (58%) |
| Past | 1,898 (48%) | 1,521 (53%) | 377 (34%) |
| Yes | 313 (7.9%) | 226 (7.9%) | 87 (7.8%) |
| **Regular physical activity** | 1,907 (48%) | 1,399 (49%) | 508 (45%) |
| **type of AF** |  |  |  |
| Paroxysmal | 1,939 (49%) | 1,318 (46%) | 621 (56%) |
| Permanent | 908 (23%) | 691 (24%) | 217 (19%) |
| Persisting | 1,111 (28%) | 832 (29%) | 279 (25%) |
| **CHAD2DS2-VASc score** | 3 (2, 4) | 3 (2, 4) | 4 (3, 5) |
| **EHRA score** |  |  |  |
| I | 1,407 (36%) | 1,098 (39%) | 309 (28%) |
| II | 773 (20%) | 516 (18%) | 257 (23%) |
| III | 189 (4.8%) | 115 (4.0%) | 74 (6.6%) |
| IV | 45 (1.1%) | 23 (0.8%) | 22 (2.0%) |
| **History of device** |  |  |  |
| None | 3,263 (82%) | 2,342 (82%) | 921 (82%) |
| CRT | 40 (1.0%) | 30 (1.1%) | 10 (0.9%) |
| CRT-ICD | 71 (1.8%) | 62 (2.2%) | 9 (0.8%) |
| ICD | 107 (2.7%) | 91 (3.2%) | 16 (1.4%) |
| Pacemaker | 455 (11%) | 305 (11%) | 150 (13%) |
| **History of pulmonal vein isolation** | 836 (21%) | 613 (22%) | 223 (20%) |
| **LVEF** | 57 (50, 61) | 55 (47, 60) | 60 (55, 65) |
| **Arterial hypertension** | 2,736 (69%) | 1,950 (69%) | 786 (70%) |
| **Diabetes mellitus** | 635 (16%) | 511 (18%) | 124 (11%) |
| **Coronary heart disease** | 1,059 (27%) | 893 (31%) | 166 (15%) |
| **Kidney disease** | 741 (19%) | 537 (19%) | 204 (18%) |
| **History of stroke/TIA** | 676 (17%) | 484 (17%) | 192 (17%) |
| **Heart failure** | 942 (24%) | 709 (25%) | 233 (21%) |
| **Beta-blocker dose %** | 12 (2, 25) | 12 (0, 25) | 12 (3, 25) |
| **RAS inhibitor dose %** | 12 (0, 50) | 14 (0, 50) | 12 (0, 40) |
| **Class IC antiarrhythmics** | 205 (5.2%) | 134 (4.7%) | 71 (6.4%) |
| **Class III Antiarrhythmics** | 725 (18%) | 521 (18%) | 204 (18%) |
| **Antiplatelet therapy** | 849 (21%) | 686 (24%) | 163 (15%) |
| **Oral anticoagulants** | 3,333 (84%) | 2,380 (84%) | 953 (85%) |

Data presented as median (interquartile range) or n(%). BMI, body mass index. AF, atrial fibrillation. EHRA, European Heart Rhythm Association. CRT, cardiac resynchronisation therapy. ICD, implantable cardioverter defibrillator. LVEF, left ventricular ejection fraction. TIA, transitory ischemic attack. RAS, renin angiotensin system. Missing data were present in BMI (n=5), heart rate (n=14), EHRA score (n=1547) and LVEF (n=2673).

### Sex-specific distribution of beta-blocker dose

For beta-blockers, the median dose prescribed was 12.5% (IQR: 1.3-25%) of the maximum dose according to the 2020 ESC guidelines for AF (4). Sixty-one (1.5%) participants were prescribed a 100% beta-blocker dose, 540 (13.7%) a 50% dose, 906 (22.9%) 25% dose, 1470 (37.0%) other doses, and 984 (24.9%) no beta-blocker. A sex-specific analysis of beta-blocker doses showed a congruent distribution across sexes in the whole population (Figure 1A), whereas patients with a history of HF showed a partial overlap between sexes (Figure 1B).

**Figure 1.**
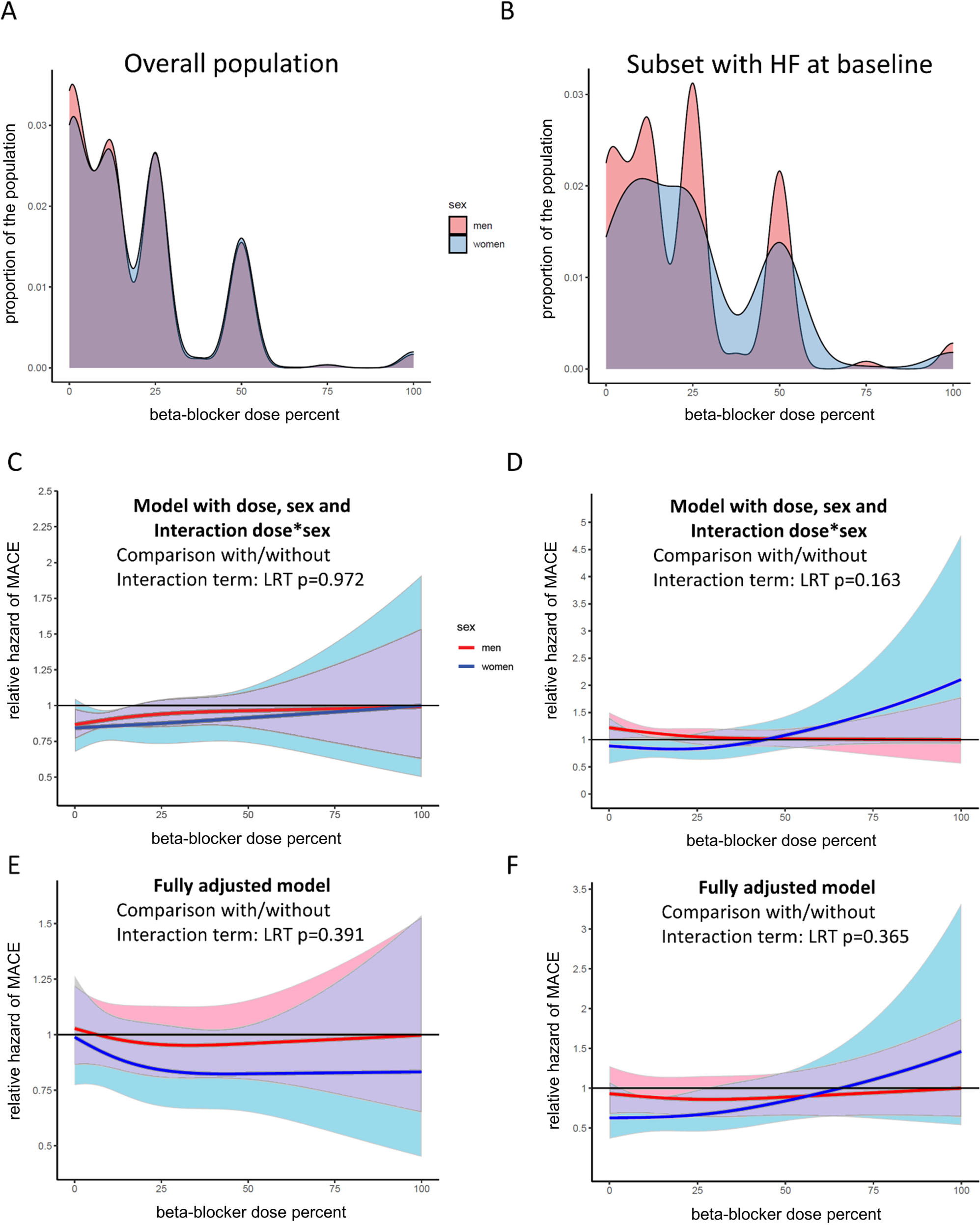
Distribution of beta-blocker dose in relation to recommended daily maximum dose according to sex in the overall population of patients with atrial fibrillation (A) and in the subset with a history of heart failure (HF) (B). Models in C-F show the hazard for MACE in the overall population (C, E) or the subset with a history of HF (D, F). Models in C and D included sex, betablocker dose and the interaction between sex and betablocker dose for the overall population (C) or the subset with a history of HF (D). Models in E and F were additionally adjusted for age, body surface area, current smoking status, regular physical activity, history of diabetes, chronic kidney disease, history of coronary artery disease, heart rate, history of hypertension, history of stroke and/or transient ischaemic attack, oral anticoagulation, antiplatelet therapy, antiarrhythmics, COPD or asthma, the dose percentage of renin angiotensin system inhibitors, and a history of HF (E only). P values of Likelihood Ratio tests (LRT) are shown to compare with models without the interaction term. MACE, major adverse cardiovascular events. Shaded areas indicate 95% confidence intervals for women (blue), men (red) or overlapping intervals (purple).

### Primary sex- specific analysis strategy of beta-blocker dose in association with MACE

As the primary strategy, we used beta-blocker drug dose in percent as a continuous variable in Cox regression models. Here, the hazard of MACE was comparable over the entire dose range of beta-blockers and for both sexes in the full population (Figure 1C) and the subgroup with a history of HF (Figure 1D). The multivariable Cox models with adjustment for clinical and demographic confounders showed a comparable hazard of MACE over the entire dose range of beta-blocker dose in the whole study population (Figure 1E) and in the subgroup with a history of HF (Figure 1F). For all these models, the interaction terms (beta-blocker dose and sex) were not significant.

### Sex-specific distribution of RAS inhibitor dose

For RAS inhibitors, the overall median dose was 12.5% (IQR: 0-50%) in the study population. Among the patients, 238 (6.0%) were prescribed a 100% RAS inhibitor dose, 590 (15.0%) a 50% dose, 502 (12.7%) a 25% dose, 984 (24.5%) other doses, and 1647 (41.8%) no RAS inhibitors. The sex-specific distribution of RAS inhibitor doses in the whole study population and the subset with pre-existing HF is shown in Figures 2A-B.

**Figure 2.**
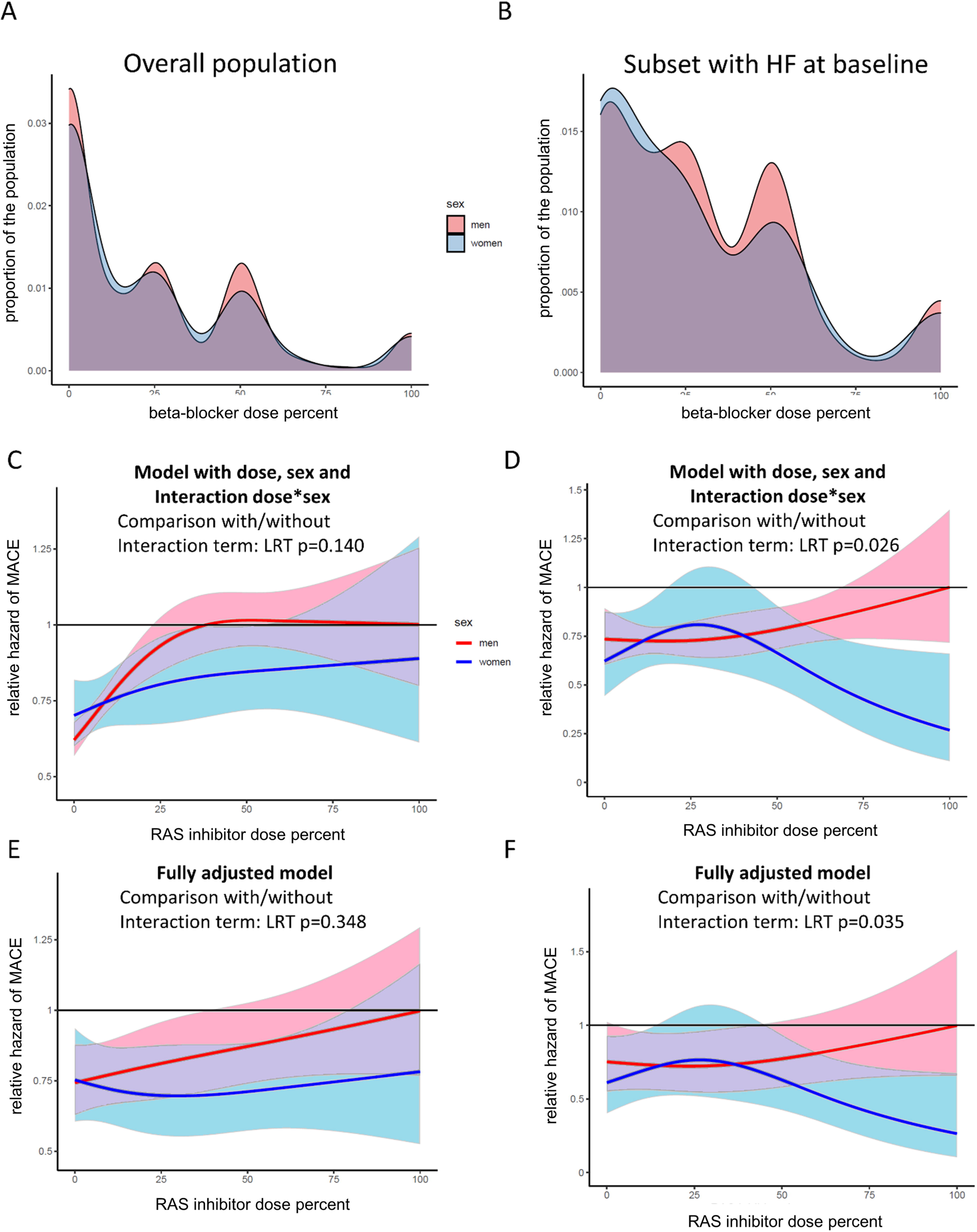
Distribution of renin angiotensin system (RAS) inhibitor dose in relation to recommended daily maximum dose according to sex in the overall population of patients with atrial fibrillation (A) and in the subset with a history of (HF) (B). Models in C-F show the hazard for MACE in the overall population (C, E) or the subset with a history of HF (D, F). Models in C and D included sex, RAS inhibitor dose and the interaction between sex and RAS inhibitor dose for the overall population (C) or the subset with a history of HF (D). Models in E and F were additionally adjusted for age, body surface area, current smoking status, regular physical activity, history of diabetes, chronic kidney disease, history of coronary artery disease, heart rate, history of hypertension, history of stroke and/or transient ischaemic attack, oral anticoagulation, antiplatelet therapy, antiarrhythmics, COPD or asthma, the dose percentage of beta-blockers, and a history of HF (E only). P values of Likelihood Ratio tests (LRT) are shown to compare with models without the interaction term. MACE, major adverse cardiovascular events. Shaded areas indicate 95% confidence intervals for women (blue), men (red) or overlapping intervals (purple).

### Primary sex- specific analysis strategy of RAS inhibitor dose in association with MACE

In the model adjusted for RAS inhibitor dose, sex and the interaction RAS inhibitor dose and sex, the hazard of MACE was lower in the RAS inhibitor dose range below 25% of drug dose in the overall study population and in the subset with HF (Figure 2C). The interaction term yielded no significant effect. However, in the subgroup with a history of HF at baseline, the hazard of MACE showed an interaction with sex (Figure 2D): In men, the hazard of MACE remained comparable across the range of RAS inhibitor doses. In women, however, an inverted u-shaped curve was present with a maximum hazard between 25% and 30% of RAS inhibitor dose and around a three-fold decline of relative hazard when approaching 100% RAS inhibitor dose. For this model, the Likelihood ratio test showed a p of 0.03 when comparing with a model not containing the interaction term of dose with sex, indicating that a relevant interaction was present. Fully adjusted models showed a similar pattern as above: The overall population showed a tendency towards a lower hazard of MACE at lower doses of RAS inhibitors (Figure 2E) without a significant interaction between dose and sex. Again, in the subgroup with a history of HF at baseline (Figure 2F), an inverted u-shaped curve was present for the hazard of MACE in women according to RAS inhibitor dose, and inclusion of the interaction term significantly affected the model (Likelihood ratio test, p=0.04).

### Secondary sex-specific analysis strategy of beta-blocker and RAS inhibitor dose in association with MACE

In the pre-specified secondary approach, we analysed drug doses of beta-blockers and RAS inhibitors as groups in quartiles together with a fifth category of those not prescribed a drug (Supplemental Figure 1). In the multivariable Cox models for the overall population and for the subgroup with a history of HF, all lower beta-blocker dose quartiles and the group prescribed no beta-blockers showed a comparable hazard of MACE in comparison to the top dose quartile, without significant interaction between beta-blocker dose group and sex (Table 2). Similar findings were made for the analyses of RAS inhibitor doses in quartiles (Table 3).

**Table 2.** Composite primary outcome according to beta-blocker dose.

| Characteristic | N | Event N | HR | 95% CI | p-value |
| --- | --- | --- | --- | --- | --- |
| **Overall population** |  |  |  |  |  |
| BB dose group | 3889 | 1123 |  |  |  |
| 4 (highest) |  |  | reference |  |  |
| 3 |  |  | 1.16 | 0.92, 1.46 | 0.20 |
| 2 |  |  | 1.11 | 0.88, 1.40 | 0.36 |
| 1 (lowest) |  |  | 1.18 | 0.94, 1.49 | 0.15 |
| 0 (no BB) |  |  | 1.18 | 0.95, 1.48 | 0.14 |
| Sex | 3889 | 1123 |  |  |  |
| Men |  |  | reference |  |  |
| Women |  |  | 0.83 | 0.60, 1.15 | 0.26 |
| BB dose group * sex | 3889 | 1123 |  |  |  |
| 3 * women |  |  | 1.1 | 0.72, 1.67 | 0.67 |
| 2 * women |  |  | 1.15 | 0.74, 1.77 | 0.53 |
| 1 * women |  |  | 1.03 | 0.67, 1.59 | 0.89 |
| 0 * women |  |  | 1.23 | 0.81, 1.86 | 0.34 |
| **Population with a history of heart failure at baseline** |  |  |  |  |  |
| BB dose group | 927 | 376 |  |  |  |
| 4 (highest) |  |  | reference |  |  |
| 3 |  |  | 0.94 | 0.65, 1.37 | 0.74 |
| 2 |  |  | 1.18 | 0.82, 1.70 | 0.37 |
| 1 (lowest) |  |  | 1.02 | 0.70, 1.49 | 0.93 |
| 0 (no BB) |  |  | 1.07 | 0.71, 1.62 | 0.75 |
| Sex | 927 | 376 |  |  |  |
| Men |  |  | reference |  |  |
| Women |  |  | 1.1 | 0.67, 1.81 | 0.69 |
| BB dose group * sex | 927 | 376 |  |  |  |
| 3 * women |  |  | 0.63 | 0.30, 1.31 | 0.22 |
| 2 * women |  |  | 0.75 | 0.37, 1.52 | 0.43 |
| 1 * women |  |  | 0.63 | 0.31, 1.30 | 0.21 |
| 0 * women |  |  | 0.64 | 0.27, 1.52 | 0.31 |

Comparison with a model not containing the interaction term of dose group \* sex: Likelihood ratio test $p=0.874$ (upper panel), $p=0.680$ (lower panel).
Among all patients, BB dose was 0% in BB dose group 0, in the range of 0.6-12.5% (minimum-maximum) in BB dose group 1, 12.5-25% in BB dose group 2, all 25% in BB dose group 03, and 25-100% in BB dose group 4. Among patients with a history of heart failure at baseline, BB dose was 0% in BB dose group 0, in the range of 1.25-12.5% in BB dose group 1, 12.5-25% in BB dose group 2, 25-50% in BB dose group 03, and 50-100% in BB dose group 4.
HR, hazard ratio. CI, confidence interval. BB, beta-blocker.

**Table 3.** Composite primary outcome according to RAS inhibitor dose.

| Characteristic | N | Event N | HR | 95% CI | p-value |
| --- | --- | --- | --- | --- | --- |
| **Overall population** |  |  |  |  |  |
| RAS inhibitor dose group | 3873 | 1113 |  |  |  |
| 4 (highest) |  |  | reference |  |  |
| 3 |  |  | 0.98 | 0.78, 1.23 | 0.85 |
| 2 |  |  | 0.83 | 0.65, 1.06 | 0.13 |
| 1 (lowest) |  |  | 0.91 | 0.72, 1.15 | 0.43 |
| 0 (no RAS inhibitor) |  |  | 0.82 | 0.66, 1.01 | 0.065 |
| Sex | 3873 | 1113 |  |  |  |
| Men |  |  | reference |  |  |
| Women |  |  | 0.87 | 0.62, 1.23 | 0.44 |
| RAS inhibitor dose group * sex | 3873 | 1113 |  |  |  |
| 3 * women |  |  | 0.85 | 0.52, 1.38 | 0.50 |
| 2 * women |  |  | 1.08 | 0.67, 1.72 | 0.76 |
| 1 * women |  |  | 1.03 | 0.65, 1.63 | 0.89 |
| 0 * women |  |  | 1.14 | 0.77, 1.68 | 0.52 |
| **Population with a history of heart failure at baseline** |  |  |  |  |  |
| RAS inhibitor dose group | 921 | 371 |  |  |  |
| 4 (highest) |  |  | reference |  |  |
| 3 |  |  | 0.94 | 0.65, 1.34 | 0.72 |
| 2 |  |  | 0.92 | 0.63, 1.34 | 0.66 |
| 1 (lowest) |  |  | 0.82 | 0.56, 1.20 | 0.30 |
| 0 (no RAS inhibitor) |  |  | 0.96 | 0.67, 1.38 | 0.83 |
| Sex | 921 | 371 |  |  |  |
| Men |  |  | reference |  |  |
| Women |  |  | 0.69 | 0.38, 1.26 | 0.23 |
| RAS inhibitor dose group * sex | 921 | 371 |  |  |  |
| 3 * women |  |  | 0.9 | 0.38, 2.16 | 0.82 |
| 2 * women |  |  | 1.16 | 0.51, 2.66 | 0.72 |
| 1 * women |  |  | 1.57 | 0.71, 3.46 | 0.26 |
| 0 * women |  |  | 1.12 | 0.53, 2.37 | 0.77 |

Comparison with a model not containing the interaction term of dose group \* sex: Likelihood ratio test p=0.729 (upper panel), p=0.725 (lower panel).
Among all patients, RAS inhibitor dose was 0% in RAS inhibitor dose group 0, in the range of 0.7-25% (minimum-maximum) in RAS inhibitor dose group 1, 25-28.6% in RAS inhibitor dose group 2, 28.6%-50% in RAS inhibitor dose group 3, and 50-100% in RAS inhibitor dose group 4.
Among patients with a history of heart failure at baseline, RAS inhibitor dose was 0% in RAS inhibitor group 0, in the range of 0.7-17.9% in RAS inhibitor group 1, 18.8-32.1% in RAS inhibitor group 2, 32.1-50% in RAS inhibitor group 3, and 50-100% in RAS inhibitor group 4.
HR, hazard ratio. CI, confidence interval. RAS, renin angiotensin system.

### Secondary outcomes

We analysed all individual components of MACE except systemic embolism as secondary outcomes using categorised beta-blocker doses (Supplemental Tables 3-6) or RAS inhibitor doses (Supplemental Tables 7-10) and their interaction with sex, respectively. The low number of events precluded an isolated assessment of systemic embolism, or an assessment of stroke in association with RAS inhibitor dose in the subgroup with a history of HF. Overall, the secondary analyses resembled those of the composite primary outcome, except for the following: The overall hazard of all-cause mortality was 0.56 (95% CI: 0.35, 0.89, p=0.01) in women compared to men (Supplemental Table 3). The third quartile of beta-blocker doses was associated with a higher hazard of stroke compared to the top quartile, with HR: 1.89 (95% CI: 1.08, 3.30; p=0.03) (Supplemental Table 4). Patients not treated with RAS inhibitors had a lower hazard of myocardial infarction compared to the top dose quartile, with HR: 0.50 (95% CI: 0.27, 0.92; p=0.03) (Supplemental Table 9). For these analyses, the interaction terms between drug dose and sex yielded no significantly different models.

### Sensitivity analyses

We performed several pre-specified and posthoc sensitivity analyses as a robustness check of the analysis strategy. All these procedures did not cause substantial changes in the results (data not shown). The inclusion of LVEF as a variable in Cox models in the subgroup of 1288 patients with available echocardiography did not modify the results. Overall, the reported findings remained robust in sensitivity analyses.

## DISCUSSION

In this study, we assessed the associations between beta-blockers or RAS inhibitors and MACE with a focus on the interaction between sex and drug dose. We found no associations between drug dose and MACE in patients with AF, but in patients with pre-existing HF, women treated with RAS inhibitors at submaximal dose showed a higher hazard of MACE compared to those treated with maximal dose.

Our data showing no overall association between beta-blockers and MACE in patients with AF are consistent with the findings of a meta-analysis by Rienstra et al. reporting that beta-blockers have no effect on mortality or hospitalisations in patients with AF, in contrast to patients with sinus rhythm (25). Prior work has, however, rarely provided sex-disaggregated data: The landmark study by van Gelder et al. who reported comparable survival between strict or lenient ventricular rate control in AF included 66% men but reported no sex-disaggregated outcomes (26). The meta-analysis on beta-blocker efficacy in patients with AF by Rienstra et al. reported a meta-regression according to the male:female sex ratio among patients of included studies but found no association between sex ratio and reported beta-blocker efficacy in studies on AF (25). Next, nearly all dose-specific analyses of beta-blocker doses in association with cardiovascular outcomes were performed among patients with HFrEF: Campodonico et al. reported that in patients with HFrEF and AF, increasing doses of beta-blockers were associated with improved patient survival (27). However, the study of Campodonico et al. included only 16% women and reported no sex-disaggregated data (27). The present findings of the Swiss-AF and BEAT-AF population containing both HFrEF and HFpEF are in contrast to some analyses restricted to patients with HFrEF in which women showed fewer deaths or HF-related hospitalisations when prescribed submaximal doses of beta-blockers (14,15). Nevertheless, the Swiss-AF and BEAT-AF cohorts differ from these populations that had a low prevalence of AF in only 35% of women and 44% of men reported by Santema et al. (14), or of any arrhythmia in 21% of women and 25% of men reported by Bots et al. (15). Of note, the meta-analysis by Kotecha et al. showed no evidence for sex as an effect modifier of beta-blocker efficacy in patients HFrEF for several cardiovascular outcomes (28). Kotecha et al. further observed no association between sex and beta-blocker discontinuation rates, which speaks rather against women with HFrEF being relatively overdosed at standard beta-blocker doses (28).

Regarding RAS inhibitors, the clinical guidelines by the European Society of Cardiology recommend an up-titration in patients with HFrEF until maximum tolerated dose is reached, but they make no specific recommendation in patients with HF with preserved LVEF (HFpEF) or AF without HF. In case a causal effect is assumed between higher dose of RAS inhibitors and lower hazard of MACE in the primary analysis, the present data would support the use of RAS inhibitors in women with AF and HF. This finding could however be due to chance because of the small number of women with 100% dose of RAS inhibitors, especially as the secondary analysis strategy of dose quartiles yielded negative results. The finding could alternatively result from bias from unmeasured confounders such as emotional stress (30) that predisposes women to adverse cardiovascular events (31–33). In addition, women could have been inadequately underdosed by treating physician, e.g. to prevent side effects. This could for instance lead to undertreated arterial hypertension in women who have arterial hypertension as comorbidity, which is associated with a higher cardiovascular risk (34). The distribution of doses was however similar between the two sexes.

Strengths of this study include the large and well-characterised cohorts analysed, the long follow-up duration and detailed medical data available, and the pressing nature of the topic that we addressed. This study has also some limitations. First, the observational design did not allow assessing causality. Thus, despite the attempts to balance the population by multivariable adjustments and inverse probability weighting, additional unmeasured confounding may remain such as frailty, intolerance of higher dosing, or change of treatment over time e.g. because of low blood pressure. Second, few patients with available echocardiography data were included, precluding the adjustment for LVEF in primary models. However, when the subset of patients with available LVEF data was analysed in a sensitivity analysis, no major changes were noted. Next, the clinical decisions in beta-blocker dosing may be influenced by previous device implantation such as pacemakers. Finally, we focused on beta-blockers and RAS inhibitors but did not assess the sex-specific doses of other drug classes that may have influenced the findings. Future studies may take additional drug classes into consideration.

Implications of the present work include that patients with AF of both sexes appeared to show dose-independent cardiovascular outcomes when treatments with beta-blockers or RAS inhibitors were prescribed. Thus, no sex-specific beta-blocker RAS inhibitor dose reconsiderations appear beneficial in patients with AF according to the present data, in contrast to what has been shown for patients with HFrEF (14). As a research implication, it is important to consider potential differences between different and overlapping populations, such as patients with AF or HF.

## Conclusion

In conclusion, the present study reveals no overall sex differences in beta-blocker or RAS inhibitor doses nor associations between beta-blocker or RAS inhibitor doses and MACE in two cohorts of patients with AF. This study adds to the emerging knowledge on sex differences in cardiovascular pharmacotherapy and can guide clinical practice.

## Acknowledgements

The authors thank the participants and collaborator of the Swiss-AF and BEAT-AF studies.

## Data availability

Due to restrictions by the Ethical Committee, data is not publicly available. Requests to access the datasets should be directed to the corresponding author.

