## Supplemental data for "Sex-specific association of cardiovascular drug doses with adverse outcomes in atrial fibrillation"

**Supplemental Table 1: Maximum pharmacotherapy dose**

| Drug | Class | Maximum daily maintenance dose |
|---|---|---|
| Bisoprolol | Beta-blocker | 20 mg |
| Carvedilol | Beta-blocker | 100 mg |
| Metoprolol XR | Beta-blocker | 400 mg |
| Nebivolol | Beta-blocker | 10 mg |
| Atenolol | Beta-blocker | 100 mg |
| Captopril | ACE inhibitor | 150 mg |
| Enalapril | ACE inhibitor | 40 mg |
| Lisinopril | ACE inhibitor | 35 mg |
| Perindopril | ACE inhibitor | 10 mg |
| Ramipril | ACE inhibitor | 10 mg |
| Trandolapril | ACE inhibitor | 4 mg |
| Candesartan | Angiotensin receptor blocker | 32 mg |
| Valsartan | Angiotensin receptor blocker | 320 mg |
| Losartan | Angiotensin receptor blocker | 150 mg |
| Irbesartan | Angiotensin receptor blocker | 300 mg |

ACE, angiotensin converting enzyme.

**Supplemental Table 2: Population with history of heart failure at baseline**

| | Overall | Men | Women |
|---|---|---|---|
| | **942** | **711** | **233** |
| **Age (years)** | 75 (68, 80) | 74 (68, 79) | 77 (70, 81) |
| **BMI (kg/m2)** | 27.3 (24.4, 30.8) | 27.5 (24.7, 30.9) | 26.7 (23.0, 30.6) |
| **BSA (m2)** | 1.94 (1.81, 2.09) | 2.00 (1.88, 2.12) | 1.76 (1.64, 1.88) |
| **Heart rate (min-1)** | 69 (60, 80) | 69 (59, 80) | 69 (60, 82) |
| **Smoking status** | | | |
| No | 367 (39%) | 241 (34%) | 126 (54%) |
| Past | 500 (53%) | 411 (58%) | 89 (38%) |
| Yes | 71 (7.5%) | 55 (7.8%) | 16 (6.9%) |
| **Regular physical activity** | 362 (38%) | 278 (39%) | 84 (36%) |
| **Type of AF** | 309 (33%) | 217 (31%) | 92 (39%) |
| Paroxysmal | 324 (34%) | 255 (36%) | 69 (30%) |
| Permanent | 309 (33%) | 237 (33%) | 72 (31%) |
| Persisting | | | |
| **CHAD2DS2-VASc score** | 5 (3, 5.75) | 3 (4, 5) | 4 (5, 6) |
| **EHRA score** | | | |
| I | 348 (37%) | 275 (39%) | 73 (31%) |
| II | 198 (21%) | 148 (21%) | 50 (21%) |
| III | 63 (6.7%) | 42 (5.9%) | 21 (9.0%) |
| IV | 18 (1.9%) | 11 (1.6%) | 7 (3.0%) |
| **History of device** | | | |
| None | 644 (68%) | 484 (68%) | 160 (69%) |
| CRT | 31 (3.3%) | 23 (3.2%) | 8 (3.4%) |
| CRT-ICD | 53 (5.6%) | 47 (6.6%) | 6 (2.6%) |
| ICD | 67 (7.1%) | 55 (7.8%) | 12 (5.2%) |
| Pacemaker | 145 (15%) | 99 (14%) | 46 (20%) |
| **History of pulmonal vein isolation** | 113 (12%) | 86 (12%) | 27 (12%) |
| **LVEF %** | 45 (35, 55) | 45 (35, 54) | 55 (38, 60) |
| **Arterial hypertension** | 751 (80%) | 566 (80%) | 185 (79%) |
| **Diabetes mellitus** | 241 (26%) | 202 (28%) | 39 (17%) |
| **Coronary heart disease** | 406 (43%) | 353 (50%) | 53 (23%) |
| **Kidney disease** | 364 (39%) | 274 (39%) | 90 (39%) |
| **History of stroke/TIA** | 169 (18%) | 130 (18%) | 39 (17%) |
| **Beta-blocker dose %** | 25 (6, 25) | 19 (6, 25) | 25 (12, 50) |
| **RAS inhibitor dose %** | 25 (3, 50) | 25 (7, 50) | 25 (0, 50) |
| **Class IC antiarrhythmics** | 5 (0.5%) | 2 (0.3%) | 3 (1.3%) |
| **Class III antiarrhythmics** | 233 (25%) | 180 (25%) | 53 (23%) |
| **Antiplatelet therapy** | 237 (25%) | 202 (28%) | 35 (15%) |
| **Oral anticoagulants** | 867 (92%) | 650 (92%) | 217 (93%) |

Data presented as median (interquartile range) or n(%). BMI, body mass index. AF, atrial fibrillation. EHRA, European Heart Rhythm Association. CRT, cardiac resynchronisation therapy. ICD, implantable cardioverter defibrillator. LVEF, left ventricular ejection fraction. TIA, transitory ischemic attack. RAS, renin angiotensin system. Missing data were present in heart rate (n=6), EHRA score (n=316) and LVEF (n=645).

**Supplemental Table 3. All-cause mortality according to BB dose**

| Characteristic | N | Event N | HR | 95% CI | p-value |
|---|---|---|---|---|---|
| **Overall population** | | | | | |
| BB dose group | 3889 | 632 | | | |
| 4 (highest) | | | reference | | |
| 3 | | | 1.05 | 0.78, 1.43 | 0.74 |
| 2 | | | 1.16 | 0.86, 1.57 | 0.32 |
| 1 (lowest) | | | 1.19 | 0.88, 1.60 | 0.25 |
| 0 (no BB) | | | 1.31 | 0.98, 1.74 | 0.07 |
| Sex | 3889 | 632 | | | |
| Men | | | reference | | |
| Women | | | 0.56 | 0.35, 0.89 | 0.01 |
| BB dose group * sex | 3889 | 632 | | | |
| 3 * women | | | 1.26 | 0.69, 2.32 | 0.45 |
| 2 * women | | | 0.96 | 0.51, 1.81 | 0.91 |
| 1 * women | | | 1.12 | 0.61, 2.06 | 0.71 |
| 0 * women | | | 1.33 | 0.74, 2.38 | 0.34 |
| **Population with a history of heart failure at baseline** | | | | | |
| BB dose group | 927 | 235 | | | |
| 4 (highest) | | | reference | | |
| 3 | | | 0.68 | 0.42, 1.09 | 0.11 |
| 2 | | | 1.19 | 0.77, 1.83 | 0.44 |
| 1 (lowest) | | | 0.87 | 0.54, 1.38 | 0.54 |
| 0 (no BB) | | | 0.99 | 0.60, 1.63 | 0.98 |
| Sex | 927 | 235 | | | |
| Men | | | reference | | |
| Women | | | 0.58 | 0.29, 1.13 | 0.11 |
| BB dose group * sex | 927 | 235 | | | |
| 3 * women | | | 0.75 | 0.26, 2.16 | 0.60 |
| 2 * women | | | 0.46 | 0.16, 1.35 | 0.16 |
| 1 * women | | | 0.8 | 0.30, 2.13 | 0.66 |
| 0 * women | | | 0.68 | 0.20, 2.35 | 0.54 |

Comparison with a model not containing the interaction term of dose group * sex: Likelihood ratio test p=0.781 (upper panel), p=0.705 (lower panel).

Among all patients, BB dose was 0% in BB dose group 0, in the range of 0.6-12.5% (minimum-maximum) in BB dose group 1, 12.5-25% in BB dose group 2, all 25% in BB dose group 03, and 25-100% in BB dose group 4.

Among patients with a history of heart failure at baseline, BB dose was 0% in BB dose group 0, in the range of 1.25-12.5% in BB dose group 1, 12.5-25% in BB dose group 2, 25-50% in BB dose group 03, and 50-100% in BB dose group 4.

Models were adjusted for age, body surface area, current smoking status, regular physical activity, history of diabetes, chronic kidney disease, history of coronary artery disease, heart rate, history of hypertension, history of stroke and/or transient ischaemic attack, oral anticoagulation, antiplatelet therapy, antiarrhythmics, COPD or asthma, the dose percentage of renin angiotensin system inhibitors, and a history of heart failure at baseline (upper panel only).

HR, hazard ratio. CI, confidence interal. BB, betablocker.

**Supplemental Table 4. Strokes according to BB dose**

| Characteristic | N | Event N | HR | 95% CI | p-value |
|---|---|---|---|---|---|
| **Overall population** | | | | | |
| BB dose group | 3889 | 189 | | | |
| 4 (highest) | | | reference | | |
| 3 | | | 1.89 | 1.08, 3.30 | 0.03 |
| 2 | | | 1.09 | 0.58, 2.03 | 0.79 |
| 1 (lowest) | | | 1 | 0.53, 1.89 | >0.99 |
| 0 (no BB) | | | 1.32 | 0.74, 2.37 | 0.35 |
| Sex | 3889 | 189 | | | |
| Men | | | reference | | |
| Women | | | 1.22 | 0.57, 2.60 | 0.61 |
| BB dose group * sex | 3889 | 189 | | | |
| 3 * women | | | 0.72 | 0.28, 1.84 | 0.50 |
| 2 * women | | | 0.66 | 0.22, 1.96 | 0.45 |
| 1 * women | | | 0.89 | 0.31, 2.55 | 0.83 |
| 0 * women | | | 1.01 | 0.39, 2.63 | 0.98 |
| **Population with a history of heart failure at baseline** | | | | | |
| BB dose group | 927 | 54 | | | |
| 4 (highest) | | | reference | | |
| 3 | | | 0.84 | 0.32, 2.24 | 0.73 |
| 2 | | | 1.16 | 0.45, 2.97 | 0.75 |
| 1 (lowest) | | | 0.5 | 0.17, 1.54 | 0.23 |
| 0 (no BB) | | | 0.71 | 0.22, 2.26 | 0.56 |
| Sex | 927 | 54 | | | |
| Men | | | reference | | |
| Women | | | 1.59 | 0.52, 4.90 | 0.42 |
| BB dose group * sex | 927 | 54 | | | |
| 3 * women | | | 0.18 | 0.02, 1.82 | 0.14 |
| 2 * women | | | 0.42 | 0.08, 2.28 | 0.32 |
| 1 * women | | | 0.72 | 0.12, 4.20 | 0.71 |
| 0 * women | | | 0.39 | 0.04, 4.39 | 0.45 |

Comparison with a model not containing the interaction term of dose group * sex: Likelihood ratio test p=0.877 (upper panel), p=0.539 (lower panel).

Among all patients, BB dose was 0% in BB dose group 0, in the range of 0.6-12.5% (minimum-maximum) in BB dose group 1, 12.5-25% in BB dose group 2, all 25% in BB dose group 03, and 25-100% in BB dose group 4.

Among patients with a history of heart failure at baseline, BB dose was 0% in BB dose group 0, in the range of 1.25-12.5% in BB dose group 1, 12.5-25% in BB dose group 2, 25-50% in BB dose group 03, and 50-100% in BB dose group 4.

Models were adjusted for age, body surface area, current smoking status, regular physical activity, history of diabetes, chronic kidney disease, history of coronary artery disease, heart rate, history of hypertension, history of stroke and/or transient ischaemic attack, oral anticoagulation, antiplatelet therapy, antiarrhythmics, COPD or asthma, the dose percentage of renin angiotensin system inhibitors, and a history of heart failure at baseline (upper panel only).

HR, hazard ratio. CI, confidence interal. BB, betablocker.

**Supplemental Table 5. Myocardial infarctions according to BB dose**

| Characteristic | N | Event N | HR | 95% CI | p-value |
|---|---|---|---|---|---|
| **Overall population** | | | | | |
| BB dose group | 3889 | 137 | | | |
| 4 (highest) | | | reference | | |
| 3 | | | 0.82 | 0.44, 1.52 | 0.53 |
| 2 | | | 0.82 | 0.44, 1.53 | 0.54 |
| 1 (lowest) | | | 0.98 | 0.54, 1.78 | 0.95 |
| 0 (no BB) | | | 0.56 | 0.28, 1.10 | 0.09 |
| Sex | 3889 | 137 | | | |
| Men | | | reference | | |
| Women | | | 0.59 | 0.23, 1.49 | 0.26 |
| BB dose group * sex | 3889 | 137 | | | |
| 3 * women | | | 1.47 | 0.42, 5.16 | 0.54 |
| 2 * women | | | 3.32 | 1.06, 10.5 | 0.04 |
| 1 * women | | | 1.67 | 0.50, 5.63 | 0.40 |
| 0 * women | | | 2.59 | 0.72, 9.30 | 0.14 |
| **Population with a history of heart failure at baseline** | | | | | |
| BB dose group | 927 | 40 | | | |
| 4 (highest) | | | reference | | |
| 3 | | | 1.05 | 0.37, 2.97 | 0.93 |
| 2 | | | 1.14 | 0.38, 3.36 | 0.82 |
| 1 (lowest) | | | 1.2 | 0.40, 3.60 | 0.75 |
| 0 (no BB) | | | 0.5 | 0.10, 2.49 | 0.40 |
| Sex | 927 | 40 | | | |
| Men | | | reference | | |
| Women | | | 1.5 | 0.41, 5.55 | 0.54 |
| BB dose group * sex | 927 | 40 | | | |
| 3 * women | | | 0.29 | 0.03, 3.32 | 0.32 |
| 2 * women | | | 1.61 | 0.27, 9.41 | 0.60 |
| 1 * women | | | 0 | 0.00, Inf | >0.99 |
| 0 * women | | | 0 | 0.00, Inf | >0.99 |

Comparison with a model not containing the interaction term of dose group * sex: Likelihood ratio test p=0.256 (upper panel), p=0.104 (lower panel).

Among all patients, BB dose was 0% in BB dose group 0, in the range of 0.6-12.5% (minimum-maximum) in BB dose group 1, 12.5-25% in BB dose group 2, all 25% in BB dose group 03, and 25-100% in BB dose group 4.

Among patients with a history of heart failure at baseline, BB dose was 0% in BB dose group 0, in the range of 1.25-12.5% in BB dose group 1, 12.5-25% in BB dose group 2, 25-50% in BB dose group 03, and 50-100% in BB dose group 4.

Models were adjusted for age, body surface area, current smoking status, regular physical activity, history of diabetes, chronic kidney disease, history of coronary artery disease, heart rate, history of hypertension, history of stroke and/or transient ischaemic attack, oral anticoagulation, antiplatelet therapy, antiarrhythmics, COPD or asthma, the dose percentage of renin angiotensin system inhibitors, and a history of heart failure at baseline (upper panel only).

HR, hazard ratio. CI, confidence interal. BB, betablocker.

**Supplemental Table 6. Hospitalisation for heart failure according to BB dose**

| Characteristic | N | Event N | HR | 95% CI | p-value |
|---|---|---|---|---|---|
| **Overall population** | | | | | |
| BB dose group | 3889 | 584 | | | |
| 4 (highest) | | | reference | | |
| 3 | | | 1.13 | 0.81, 1.56 | 0.47 |
| 2 | | | 1.27 | 0.93, 1.75 | 0.14 |
| 1 (lowest) | | | 1.32 | 0.96, 1.80 | 0.09 |
| 0 (no BB) | | | 1.16 | 0.84, 1.59 | 0.36 |
| Sex | 3889 | 584 | | | |
| Men | | | reference | | |
| Women | | | 1.08 | 0.70, 1.65 | 0.74 |
| BB dose group * sex | 3889 | 584 | | | |
| 3 * women | | | 1.1 | 0.63, 1.93 | 0.74 |
| 2 * women | | | 0.82 | 0.46, 1.48 | 0.52 |
| 1 * women | | | 0.9 | 0.51, 1.59 | 0.72 |
| 0 * women | | | 0.9 | 0.50, 1.62 | 0.74 |
| **Population with a history of heart failure at baseline** | | | | | |
| BB dose group | 927 | 218 | | | |
| 4 (highest) | | | reference | | |
| 3 | | | 1.01 | 0.60, 1.70 | 0.96 |
| 2 | | | 1.3 | 0.79, 2.14 | 0.30 |
| 1 (lowest) | | | 1.13 | 0.68, 1.89 | 0.63 |
| 0 (no BB) | | | 0.96 | 0.54, 1.72 | 0.90 |
| Sex | 927 | 218 | | | |
| Men | | | reference | | |
| Women | | | 1.48 | 0.78, 2.78 | 0.23 |
| BB dose group * sex | 927 | 218 | | | |
| 3 * women | | | 0.73 | 0.29, 1.82 | 0.50 |
| 2 * women | | | 0.63 | 0.26, 1.51 | 0.30 |
| 1 * women | | | 0.53 | 0.21, 1.32 | 0.17 |
| 0 * women | | | 0.68 | 0.23, 2.04 | 0.49 |

Comparison with a model not containing the interaction term of dose group * sex: Likelihood ratio test p=0.879 (upper panel), p=0.719 (lower panel).

Among all patients, BB dose was 0% in BB dose group 0, in the range of 0.6-12.5% (minimum-maximum) in BB dose group 1, 12.5-25% in BB dose group 2, all 25% in BB dose group 03, and 25-100% in BB dose group 4.

Among patients with a history of heart failure at baseline, BB dose was 0% in BB dose group 0, in the range of 1.25-12.5% in BB dose group 1, 12.5-25% in BB dose group 2, 25-50% in BB dose group 03, and 50-100% in BB dose group 4.

Models were adjusted for age, body surface area, current smoking status, regular physical activity, history of diabetes, chronic kidney disease, history of coronary artery disease, heart rate, history of hypertension, history of stroke and/or transient ischaemic attack, oral anticoagulation, antiplatelet therapy, antiarrhythmics, COPD or asthma, the dose percentage of renin angiotensin system inhibitors, and a history of heart failure at baseline (upper panel only).

HR, hazard ratio. CI, confidence interal. BB, betablocker.

**Supplemental Table 7. All-cause mortality according to RAS inhibitor dose**

| Characteristic | N | Event N | HR | 95% CI | p-value |
|---|---|---|---|---|---|
| **Overall population** | | | | | |
| RAS inhibitor dose group | 3873 | 630 | | | |
| 4 (highest) | | | reference | | |
| 3 | | | 1.01 | 0.75, 1.36 | 0.94 |
| 2 | | | 0.79 | 0.57, 1.09 | 0.15 |
| 1 (lowest) | | | 0.87 | 0.64, 1.18 | 0.38 |
| 0 (no RAS inhibitor) | | | 0.83 | 0.64, 1.10 | 0.19 |
| Sex | 3873 | 630 | | | |
| Men | | | reference | | |
| Women | | | 0.67 | 0.42, 1.06 | 0.09 |
| RAS inhibitor dose group * sex | 3873 | 630 | | | |
| 3 * women | | | 0.75 | 0.38, 1.46 | 0.39 |
| 2 * women | | | 1.11 | 0.58, 2.11 | 0.76 |
| 1 * women | | | 1.01 | 0.54, 1.88 | 0.98 |
| 0 * women | | | 0.92 | 0.53, 1.58 | 0.76 |
| **Population with a history of heart failure at baseline** | | | | | |
| RAS inhibitor dose group | 921 | 235 | | | |
| 4 (highest) | | | reference | | |
| 3 | | | 0.96 | 0.61, 1.49 | 0.84 |
| 2 | | | 0.96 | 0.61, 1.52 | 0.87 |
| 1 (lowest) | | | 0.75 | 0.47, 1.20 | 0.23 |
| 0 (no RAS inhibitor) | | | 0.98 | 0.64, 1.52 | 0.94 |
| Sex | 921 | 235 | | | |
| Men | | | reference | | |
| Women | | | 0.35 | 0.15, 0.84 | 0.02 |
| RAS inhibitor dose group * sex | 921 | 235 | | | |
| 3 * women | | | 0.92 | 0.26, 3.28 | 0.90 |
| 2 * women | | | 1.06 | 0.32, 3.48 | 0.92 |
| 1 * women | | | 1.82 | 0.58, 5.69 | 0.30 |
| 0 * women | | | 1.16 | 0.39, 3.44 | 0.79 |

Comparison with a model not containing the interaction term of dose group * sex: Likelihood ratio test p=0.830 (upper panel), p=0.786 (lower panel).
Among all patients, RAS inhibitor dose was 0% in RAS inhibitor dose group 0, in the range of 0.7-25% (minimum-maximum) in RAS inhibitor dose group 1, 25-28.6% in RAS inhibitor dose group 2, 28.6%-50% in RAS inhibitor dose group 3, and 50-100% in RAS inhibitor dose group 4.
Among patients with a history of heart failure at baseline, RAS inhibitor dose was 0% in RAS inhibitor dose group 0, in the range of 0.7-17.9% in RAS inhibitor dose group 1, 18.8-32.1% in RAS inhibitor dose group 2, 32.1-50% in RAS inhibitor dose group 3, and 50-100% in RAS inhibitor dose group 4.
Models were adjusted for age, body surface area, current smoking status, regular physical activity, history of diabetes, chronic kidney disease, history of coronary artery disease, heart rate, history of hypertension, history of stroke and/or transient ischaemic attack, oral anticoagulation, antiplatelet therapy, antiarrhythmics, COPD or asthma, the dose percentage of beta-blockers, and a history of heart failure at baseline (upper panel only).

HR, hazard ratio. CI, confidence interal. RAS, renin angiotensin system.

**Supplemental Table 8. Strokes according to RAS inhibitor dose**

| Characteristic | N | Event N | HR | 95% CI | p-value |
|---|---|---|---|---|---|
| **Overall population** | | | | | |
| RAS inhibitor dose group | 3873 | 187 | | | |
| 4 (highest) | | | reference | | |
| 3 | | | 1.09 | 0.59, 2.02 | 0.79 |
| 2 | | | 0.81 | 0.41, 1.61 | 0.55 |
| 1 (lowest) | | | 1.04 | 0.55, 1.95 | 0.91 |
| 0 (no RAS inhibitor) | | | 1.14 | 0.66, 1.95 | 0.64 |
| Sex | 3873 | 187 | | | |
| Men | | | reference | | |
| Women | | | 1.11 | 0.48, 2.55 | 0.81 |
| RAS inhibitor dose group * sex | 3873 | 187 | | | |
| 3 * women | | | 0.53 | 0.15, 1.86 | 0.32 |
| 2 * women | | | 0.88 | 0.26, 2.93 | 0.84 |
| 1 * women | | | 0.76 | 0.24, 2.40 | 0.64 |
| 0 * women | | | 1.15 | 0.46, 2.86 | 0.77 |

Comparison with a model not containing the interaction term of dose group * sex: Likelihood ratio test p=0.639 (upper panel), p=0.028 (lower panel).
Among all patients, RAS inhibitor dose was 0% in RAS inhibitor dose group 0, in the range of 0.7-25% (minimum-maximum) in RAS inhibitor dose group 1, 25-28.6% in RAS inhibitor dose group 2, 28.6%-50% in RAS inhibitor dose group 3, and 50-100% in RAS inhibitor dose group 4.
Models were adjusted for age, body surface area, current smoking status, regular physical activity, history of diabetes, chronic kidney disease, history of coronary artery disease, heart rate, history of hypertension, history of stroke and/or transient ischaemic attack, oral anticoagulation, antiplatelet therapy, antiarrhythmics, COPD or asthma, the dose percentage of beta-blockers, and a history of heart failure at baseline.
HR, hazard ratio. CI, confidence interal. RAS, renin angiotensin system.

**Supplemental Table 9. Myocardial infarctions according to RAS inhibitor dose**

| Characteristic | N | Event N | HR | 95% CI | p-value |
|---|---|---|---|---|---|
| **Overall population** | | | | | |
| RAS inhibitor dose group | 3873 | 134 | | | |
| 4 (highest) | | | reference | | |
| 3 | | | 0.75 | 0.39, 1.44 | 0.39 |
| 2 | | | 0.79 | 0.40, 1.53 | 0.48 |
| 1 (lowest) | | | 0.91 | 0.48, 1.73 | 0.78 |
| 0 (no RAS inhibitor) | | | 0.5 | 0.27, 0.92 | 0.026 |
| Sex | 3873 | 134 | | | |
| Men | | | reference | | |
| Women | | | 1.41 | 0.63, 3.15 | 0.40 |
| RAS inhibitor dose group * sex | 3873 | 134 | | | |
| 3 * women | | | 0.57 | 0.15, 2.14 | 0.40 |
| 2 * women | | | 0.61 | 0.17, 2.15 | 0.44 |
| 1 * women | | | 0.5 | 0.14, 1.73 | 0.27 |
| 0 * women | | | 1.2 | 0.45, 3.22 | 0.71 |
| **Population with a history of heart failure at baseline** | | | | | |
| RAS inhibitor dose group | 921 | 38 | | | |
| 4 (highest) | | | reference | | |
| 3 | | | 0.6 | 0.19, 1.86 | 0.38 |
| 2 | | | 0.75 | 0.24, 2.36 | 0.62 |
| 1 (lowest) | | | 0.56 | 0.17, 1.89 | 0.35 |
| 0 (no RAS inhibitor) | | | 0.78 | 0.27, 2.27 | 0.65 |
| Sex | 921 | 38 | | | |
| Men | | | reference | | |
| Women | | | 2.2 | 0.59, 8.18 | 0.24 |
| RAS inhibitor dose group * sex | 921 | 38 | | | |
| 3 * women | | | 0.79 | 0.10, 6.26 | 0.83 |
| 2 * women | | | 0.24 | 0.02, 2.91 | 0.26 |
| 1 * women | | | 0.32 | 0.03, 4.09 | 0.38 |
| 0 * women | | | 0.16 | 0.01, 1.96 | 0.15 |

Comparison with a model not containing the interaction term of dose group * sex: Likelihood ratio test p=0.471 (upper panel), p=0.499 (lower panel).

Among all patients, RAS inhibitor dose was 0% in RAS inhibitor dose group 0, in the range of 0.7-25% (minimum-maximum) in RAS inhibitor dose group 1, 25-28.6% in RAS inhibitor dose group 2, 28.6%-50% in RAS inhibitor dose group 3, and 50-100% in RAS inhibitor dose group 4.

Among patients with a history of heart failure at baseline, RAS inhibitor dose was 0% in RAS inhibitor dose group 0, in the range of 0.7-17.9% in RAS inhibitor dose group 1, 18.8-32.1% in RAS inhibitor dose group 2, 32.1-50% in RAS inhibitor dose group 3, and 50-100% in RAS inhibitor dose group 4.

Models were adjusted for age, body surface area, current smoking status, regular physical activity, history of diabetes, chronic kidney disease, history of coronary artery disease, heart rate, history of hypertension, history of stroke and/or transient ischaemic attack, oral anticoagulation, antiplatelet therapy, antiarrhythmics, COPD or asthma, the dose percentage of beta-blockers, and a history of heart failure at baseline (upper panel only).

HR, hazard ratio. CI, confidence interal. RAS, renin angiotensin system.

**Supplemental Table 10. Hospitalisation for heart failure according to RAS inhibitor dose**

| Characteristic | N | Event N | HR | 95% CI | p-value |
|---|---|---|---|---|---|
| **Overall population** | | | | | |
| RAS inhibitor dose group | 3873 | 579 | | | |
| 4 (highest) | | | reference | | |
| 3 | | | 1.04 | 0.76, 1.43 | 0.79 |
| 2 | | | 0.8 | 0.56, 1.13 | 0.21 |
| 1 (lowest) | | | 0.95 | 0.69, 1.31 | 0.74 |
| 0 (no RAS inhibitor) | | | 0.81 | 0.60, 1.08 | 0.16 |
| Sex | 3873 | 579 | | | |
| Men | | | reference | | |
| Women | | | 0.93 | 0.58, 1.48 | 0.75 |
| RAS inhibitor dose group * sex | 3873 | 579 | | | |
| 3 * women | | | 0.92 | 0.49, 1.76 | 0.81 |
| 2 * women | | | 1.27 | 0.67, 2.40 | 0.47 |
| 1 * women | | | 1.13 | 0.61, 2.10 | 0.69 |
| 0 * women | | | 1.11 | 0.65, 1.92 | 0.70 |
| **Population with a history of heart failure at baseline** | | | | | |
| RAS inhibitor dose group | 921 | 216 | | | |
| 4 (highest) | | | reference | | |
| 3 | | | 1.02 | 0.63, 1.66 | 0.93 |
| 2 | | | 0.96 | 0.57, 1.61 | 0.87 |
| 1 (lowest) | | | 0.97 | 0.59, 1.59 | 0.89 |
| 0 (no RAS inhibitor) | | | 0.96 | 0.59, 1.58 | 0.88 |
| Sex | 921 | 216 | | | |
| Men | | | reference | | |
| Women | | | 1.18 | 0.59, 2.37 | 0.63 |
| RAS inhibitor dose group * sex | 921 | 216 | | | |
| 3 * women | | | 0.79 | 0.29, 2.15 | 0.65 |
| 2 * women | | | 0.98 | 0.37, 2.56 | 0.96 |
| 1 * women | | | 0.94 | 0.36, 2.42 | 0.89 |
| 0 * women | | | 0.65 | 0.26, 1.65 | 0.37 |

Comparison with a model not containing the interaction term of dose group * sex: Likelihood ratio test p=0.892 (upper panel), p=0.888 (lower panel).
Among all patients, RAS inhibitor dose was 0% in RAS inhibitor dose group 0, in the range of 0.7-25% (minimum-maximum) in RAS inhibitor dose group 1, 25-28.6% in RAS inhibitor dose group 2, 28.6%-50% in RAS inhibitor dose group 3, and 50-100% in RAS inhibitor dose group 4.
Among patients with a history of heart failure at baseline, RAS inhibitor dose was 0% in RAS inhibitor dose group 0, in the range of 0.7-17.9% in RAS inhibitor dose group 1, 18.8-32.1% in RAS inhibitor dose group 2, 32.1-50% in RAS inhibitor dose group 3, and 50-100% in RAS inhibitor dose group 4.
Models were adjusted for age, body surface area, current smoking status, regular physical activity, history of diabetes, chronic kidney disease, history of coronary artery disease, heart rate, history of hypertension, history of stroke and/or transient ischaemic attack, oral anticoagulation, antiplatelet therapy, antiarrhythmics, COPD or asthma, the dose percentage of beta-blockers, and a history of heart failure at baseline (upper panel only).

HR, hazard ratio. CI, confidence interal. RAS, renin angiotensin system.

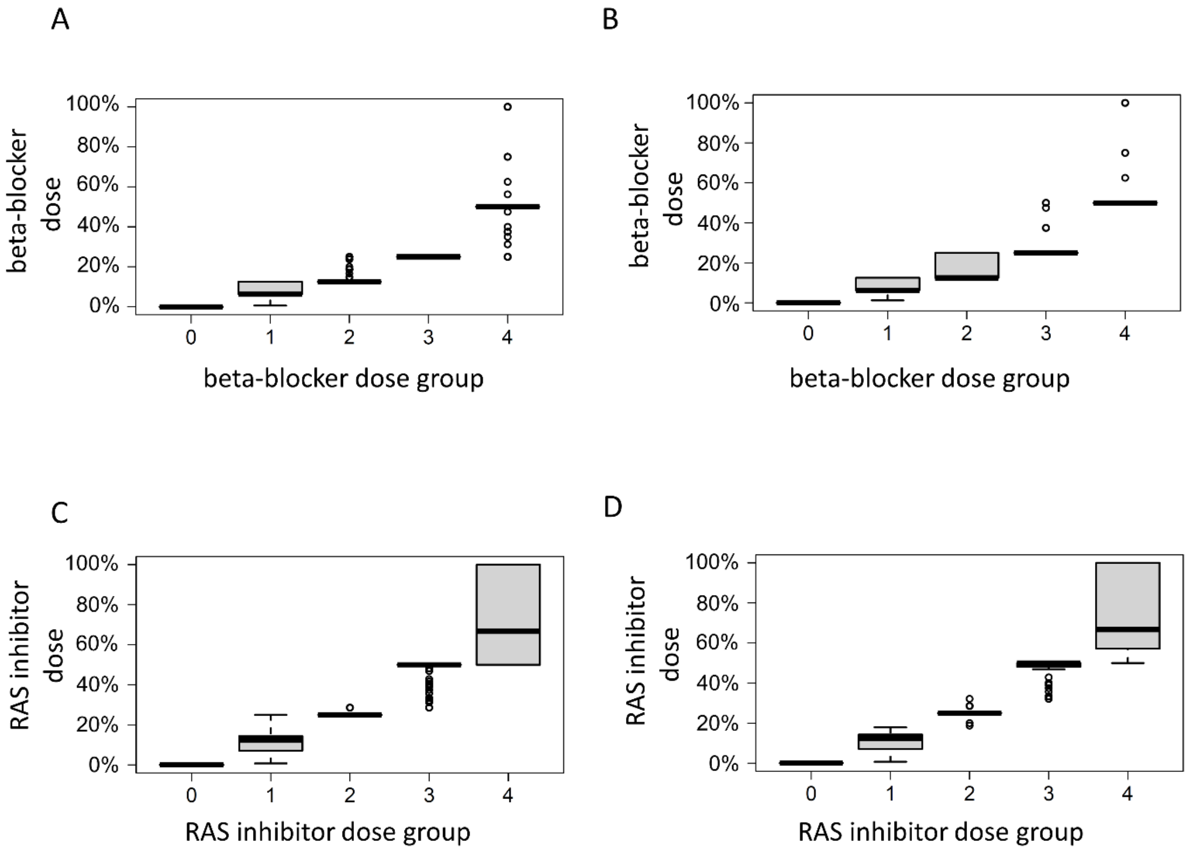


**Supplemental Figure 1.** Distribution of beta-blocker dose (A, B) and renin angiotensin system (RAS) Inhibitor dose (C-D) in percent of maximum dose in the quartiles 1-4 and those without the respective drug. Populations of all patients with atrial fibrillation (A, C) or the subset with atrial fibrillation and a history of heart failure are displayed (B, D).
